# A Seven-Year Retrospective Assessment Evaluating the Effectiveness of Electronic Consultation (e-Consult) Service in Genetics

**DOI:** 10.1101/2024.04.29.24306521

**Authors:** Sawona Biswas, Joyce So, Robert Wallerstein, Ralph Gonzales, Delphine Tout, Lisa DeAngelis, Aleksandar Rajkovic

## Abstract

Electronic consultation (e-Consult) programs serve as a conduit between healthcare providers and specialized genetic experts. This retrospective chart review and summary report presents the experience of implementing a Genetics e-Consult Service at the University of California San Francisco (UCSF) from 2016 through 2024 across multiple disciplines. The study examines 622 requests managed by the Genetics team, resulting in the completion of 360 e-Consults (57.8%) and the decline of 262 e-Consults (42.1%). Provider-to-provider consultations, conducted by board-certified geneticists, were completed within 3 days (83.9%), with consultation times ranging from 5 to 20 minutes in most cases (67%). Most requests originated from general practitioners in primary care, pediatrics, and family medicine (48.1%). Analysis of a subset of e-Consults (n=144) revealed that diagnostic queries accounted for 50.6% of requests, followed by management of symptoms (17%) and test interpretation (11%). Providers adhered to geneticists’ recommendations in 84% of cases. These findings underscore the potential of e-Consult frameworks as a viable strategy to enhance accessibility to genetic healthcare services.

## INTRODUCTION

The field of genetics and genomics has experienced significant advancements in recent years, leading to increased integration of genetic testing and considerations into various medical disciplines. However, the number of genetic subspecialists remains limited, resulting in challenges for patients and healthcare providers seeking timely access to genetics expertise^1,2^. The shortage of genetic specialists, coupled with the growing demand for their services, has led to prolonged waiting times for appointments and potential delays in patient care^3^.

As genetic testing is increasingly incorporated into the diagnostic and treatment processes across medical specialties, there is a pressing need to improve communication and collaboration between genetic specialists and a wide range of healthcare providers^4.^ Primary care clinicians, specialists in other disciplines, and allied health professionals often require guidance and support when interpreting genetic test results, understanding their implications, and supporting patient informed decisions regarding their management ^5.^

To address these challenges, healthcare systems have explored innovative solutions, such as electronic consultations (e-Consults), to facilitate prompt access to specialist expertise ^6^. E-Consults have emerged as a promising approach to bridge the gap between the limited number of genetic specialists and the growing demand for their service. Within genetics and genomics programs, e-Consult initiatives have demonstrated their effectiveness in addressing basic questions without the need for an in-person visit, and providing educational opportunities for generalists to enhance their knowledge and skills in genetics care ^7-10^. As genetics becomes increasingly integrated into mainstream medical practice, e-Consults serve as a valuable tool to foster collaboration, knowledge sharing, and the provision of high-quality, genetics-informed care to patients.

The e-Consult service at the University of California San Francisco (UCSF) started in 2012^11^. Initially, the program was grant funded and then financially supported by the health system. By 2015, UCSF entered into agreements with three payers to receive reimbursement for specialist e-Consults, and by 2020, the Centers for Medicare and Medicaid Services (CMS) and other payers started providing reimbursement for e-Consults. The introduction of Genetics e-Consults occurred in June 2017, positioning UCSF as an early adopter in the United States. Here we describe the establishment of our Genetics e-Consult service for providers within our institution and its utilization.

## METHODS

### Process of e-consultation

Ordering providers within UCSF Health can initiate an e-Consult request through our hospital’s electronic medical record (EPIC Hyperspace 2023), Providers can choose from the Adult Genetics, Pediatrics Genetics or Cancer Risk Genetics service. After selecting the service, the ordering providers will be prompted to choose one of the four diagnosis-specific SmartText templates: 1. Query genetic condition/syndrome 2. Known genetic condition/syndrome 3. Family history of genetic condition or 4. Other. Each template will guide the provider through different open-ended questions that capture the reason for e-Consult, relevant test results, family history information, and any other pertinent information. After submission, the e-Consult request is routed to the designated pool and the e-Consultant on service reviews the details and responds.

The Genetics e-Consultant summarizes or refines the consultative question, offers an individualized recommendation along with supporting evidence or current best practices, and outlines a contingency plan (including guidelines for when to refer for an in-person or telehealth specialty visit). The standard response time for an e-Consult is set at three business days. In cases where the clinical complexity exceeds the scope of an e-Consult, the e-Consultant may opt to recommend a traditional new patient appointment. The e-Consultant will cc: their specialty staff on the e-Consult response who then convert the e-Consult order to a referral order for routing to the appropriate scheduling team. (Upon submittal, requestor indicates ok to convert to referral or not ok to convert). Alternatively, if the consultation falls outside the consultant’s area of expertise and is better suited for another specialty, the e-Consultant will decline the e-Consult with that recommendation. Within our department, a dedicated team of geneticists is responsible for addressing e-Consults. Once the e-Consult is completed or declined, it is returned to the referring provider’s EPIC inbox with either the clinical recommendations or reason for decline.

### Retrospective chart review

A retrospective chart review was conducted to explore the details of e-Consults ordered from Oct 2016 to March 2024. This retrospective quality control/quality improvement (QC/QI) study involved a chart review of de-identified data from the electronic medical records at the University of California San Francisco (UCSF). The study was conducted solely for the purpose of evaluating and improving the existing Genetics e-Consult service within UCSF. As this project did not involve primary data collection and was intended for internal quality improvement with no impact on patient care decisions or patient rights, it did not constitute human subjects research under the purview of the Department of Health and Human Services. The Ethics Committee/Institutional Review Board (IRB) of University of California San Francisco (UCSF) waived ethical approval as per UCSF’s internal policies and federal regulations. Our group characterized the ordering providers, departments, referral groups, descriptions of the consult, primary referral diagnosis, consult status, response times, e-Consult recommendations, number of related referrals to Genetics and tests performed. Data was collected from EPIC reports, individual chart review, and descriptive statistical analysis was completed using R software and Microsoft XML. Statistical reporting includes all e-Consult activity originating from an order. Per UCSF Health e-Consult EPIC workflow, all e-Consult orders result in a completed and closed encounter for reporting, whether completed with a clinical recommendation or declined for the reason indicated.

## RESULT

From October 2016 to March 2024, the UCSF Genetics e-Consult service received a total of 622 consultation orders, with yearly volumes increasing from 34 in FY 2017 to 144 in FY 2023 [TABLE 1]. Out of 622 Genetics e-Consults, 360 (57.8%) were completed, while 262 (42.1%) were either declined or deferred to another specialty [TABLE 2]. Same day consults were answered in 42.3%, 83.9% of cases were answered within three days and 90.4% were answered within seven days [TABLE 3].

**Table 1:** Total number of e-Consults received by Genetics at UCSF from 10/01/2016 till 03/01/2024.

|  |  | **Total**<br>**To Date** | **FY**<br>**2024\*** | **FY**<br>**2023** | **FY**<br>**2022** | **FY**<br>**2021** | **FY**<br>**2020** | **FY**<br>**2019** | **FY**<br>**2018** | **FY**<br>**2017** |
| --- | --- | --- | --- | --- | --- | --- | --- | --- | --- | --- |
| Specialty | Start Date | Total<br>eConsult<br>Orders | Total<br>eConsult<br>Orders | Total<br>eConsult<br>Orders | Total<br>eConsult<br>Orders | Total<br>eConsult<br>Orders | Total<br>eConsult<br>Orders | Total<br>eConsult<br>Orders | Total<br>eConsult<br>Orders | Total<br>eConsult<br>Orders |
| Adult Cancer<br>Risk Genetics | 3/1/2023 | 19 | 9 | 10 |  |  |  |  |  |  |
| Adult Genetics | 7/1/2020 | 349 | 52 | 109 | 100 | 88 |  |  |  |  |
| Pediatric Genetics | 4/1/2020 | 50 | 5 | 25 | 11 | 9 |  |  |  |  |
| Pediatric and<br>Adult Genetics | 10/1/2016 | 204 |  |  |  |  | 58 | 61 | 51 | 34 |
| **TOTAL** |  | **622** | **66** | **144** | **111** | **97** | **58** | **61** | **51** | **34** |

**Table 2:** Details of the total 262 e-consults declined and reasons.

| **Genetics e-Consult Volume**<br>**(by Orders, Completed, Declined)** | **Total e-Consult**<br>**Orders** | **Total**<br>**Completed** | **Total**<br>**Declined** | **%**<br>**Declined** |
| --- | --- | --- | --- | --- |
| To Date | 622 | 360 | 262 | 42.1% |

Table 2: Details of the total 262 e-consults declined and reasons.
| **Genetics e-Consults - Decline Reasons** | **Volume of**<br>**Declines** | **% Distribution of**<br>**Decline Reasons** |
| --- | --- | --- |
| Decline and Schedule | 184 | 70.2% |
| Decline, PCP Preference not to schedule | 1 | 0.4% |
| Decline, Established patient & problem | 3 | 1.1% |
| Decline, Insufficient Info | 3 | 1.1% |
| Decline, Other | 71 | 27.1% |
| **Total** | **262** | **100.0%** |

**Table 3:** Percentage response times of the total 622 e-consults orders answered within the same day, three business days and seven business days.

| **Response Time (Goal = 3 BUSINESS days)** | **% Answered Same Day** | **% Answered within 3 Days** | **% Answered within 7 Days** |
| --- | --- | --- | --- |
| **Genetics** | 42.3% | 83.9% | 90.4% |
| **UCSF Program - All Specialties** | 37.6% | 87.9% | 95.9% |

A comprehensive case-level analysis of the 144 e-Consults ordered during fiscal year 2023 (June 2022 – July 2023) revealed that Adult Genetics received most consults (75.7%), followed by Pediatric Genetics (17.4%) and Adult Cancer Risk Genetics (6.9%) [TABLE 1]. 61.1% of these e-Consults were completed with detailed responses, 31.9% were declined and converted to in-person Genetics appointments, and 6.9% were declined for other reasons such as insufficient information or referral to another specialty. Nearly half (47.9%) of the FY 2023 e-Consults originated from general practices like Primary Care, General Medicine, and Women’s Health [TABLE 1]. e-Consultants spent 11-20 minutes on 39.1% of cases, 5-10 minutes on 28%, 21-30 minutes on 17.8%, less than 5 minutes on 7.9%, and over 31 minutes on 7.1% of cases [TABLE 4]. The most common types of e-Consult requests were queries about genetic diagnoses (50.6%), assistance with test interpretation (11.1%), and management recommendations for patients with known (9%) or without known (8.3%) genetic diagnoses [TABLE 5]. e-Consultants provided a range of services, with the most frequent being scheduling patients for outpatient visits (42.3%), providing genetic counseling and management recommendations (24.3%), making management recommendations alone (18.5%), and offering guidance on genetic testing (11.8%) [TABLE 6]. Chart review of 118 e-Consults revealed that 48% of patients were referred to and seen by Genetics, 12% were referred with a pending appointment, 4% were referred but cancelled their appointments, 20% had their care informed by the e-Consult recommendations, 4% had genetic testing ordered by their referring provider, 10% did not get a Genetics referral, and 2% did not have any genetic testing performed [FIGURE 1].

**Table 4:** Distribution of e-Consultant time spent on each consult for all 622 e-Consults.

| **Distribution of e-Consultant Time Spent** | **%** |
| --- | --- |
| Under 5 minutes | 7.9% |
| 5-10 minutes | 28.0% |
| 11-20 minutes | 39.1% |
| 21-30 minutes | 17.8% |
| 31 minutes or greater | 7.1% |
| **Total** | 100.0% |

**Table 5:** Details of the types of e-Consults requested in Fiscal Year 2023 (count and percentages).

| **Category of e-Consult request** | **Count** | **%** |
| --- | --- | --- |
| Query genetic diagnosis | 73 | 50.6% |
| Help with test interpretation | 16 | 11.1% |
| Help with management of patient(s) with known genetic diagnosis | 13 | 9.0% |
| Help with management of patient(s) without known genetic diagnosis | 12 | 8.3% |
| Help with identifying the right test order, laboratory, or how to order | 12 | 8.3% |
| Help with referral appropriateness | 7 | 4.8% |
| Family history of a genetic condition | 7 | 4.8% |
| Prenatal/Preconception counseling | 4 | 2.7% |
| **Grand Total** | **144** |  |

**Table 6:** Details of the types of action(s) or outcome(s) undertaken by the e-Consultant in fiscal year 2023 after the e-Consult is completed (count, total count per group, and percentages).

| **E-Consultant response action group** | **Count** | **Total count per group** |
| --- | --- | --- |
| Schedule for outpatient visit | 54 |  |
| Schedule for outpatient visit, and management recommendations made | 4 | 61(42.3%) |
| Schedule for outpatient visit, and interpreted test results | 1 |  |
| Schedule for outpatient visit, and provided test details with or without links | 2 |  |
| Conducted provider education, and management recommendations made | 15 |  |
| Conducted provider education, and recommended genetics referral | 7 | 35 (24.3%) |
| Conducted provider education, and recommended to test family members | 6 |  |
| Conducted provider education, and provided test details with or without links | 1 |  |
| Conducted provider education | 6 |  |
| Management recommendations made | 13 |  |
| Management recommendations made, and interpreted test results | 5 | 26 (18.5%) |
| Management recommendations made, and recommended genetics referral | 4 |  |
| Management recommendations made, provided test details with or without links | 2 |  |
| Management recommendations made, and recommended to test family members | 2 |  |
| Provided test details with or without links | 10 |  |
| Provided test details with or without links, and recommended genetics referral | 5 | 17 (11.8%) |
| Provided test details with or without links, and interpreted test results | 2 |  |
| Recommended genetics referral | 4 |  |
| Recommended genetics referral, and interpreted test results | 1 | 5 (3.4%) |
| **Grand Total** | **144** |  |

**Figure 1:**
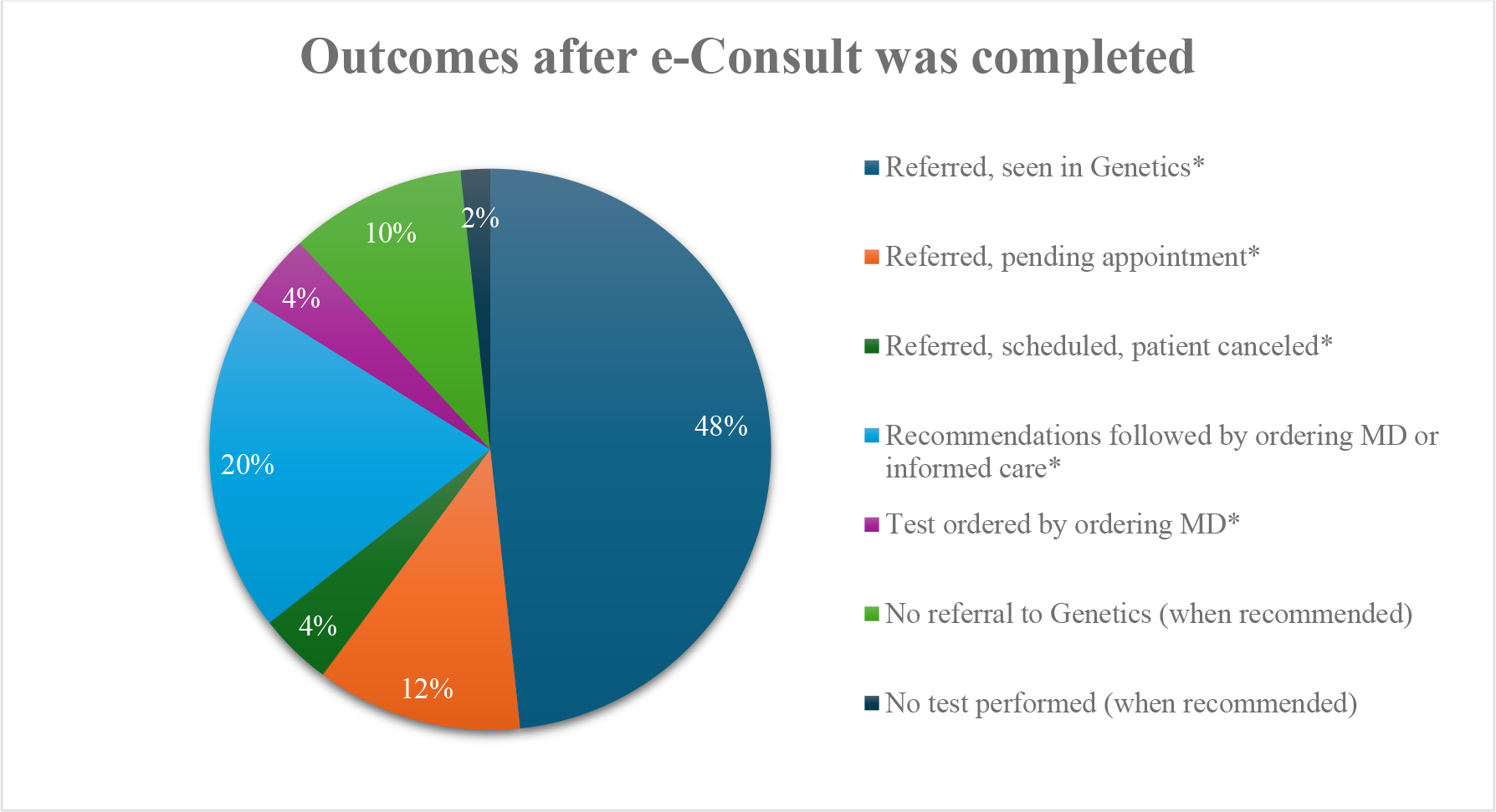
Distribution of the outcomes of the 144 e-Consults documented in patient chart. *Referred – means a referral to Genetics was initiated after the e-Consultation was completed. *Scheduled or Seen in Genetics – means the patient was scheduled for an outpatient visit with a geneticist or a genetic counselor. *Recommendations – means any tests, treatments suggestions, actions, or opinions shared by the e-Consultant with the ordering provider. *Informed care – means that the ordering provider, after receiving recommendations from the e-Consultant, changed management or informed care.

## DISCUSSION

Our study demonstrates the successful implementation and growth of an electronic consultation (e-Consult) service connecting general providers and specialists with genetics experts at a large academic medical center. The UCSF Genetics e-Consult service experienced a steady increase in volume from 34 consults in FY 2017 to 144 in FY 2023 [TABLE 1], highlighting the growing demand for accessible genetics expertise.

The e-Consult service provided an efficient means to triage genetics questions, with 57.8% of the 622 submitted consults completed and 42.1% declined, primarily to schedule in-person evaluations instead (70.2% of declined cases) [TABLE 2]. The average turnaround time of 3 days demonstrates the responsiveness of the service in addressing clinical questions.

Analysis of 144 e-Consults from FY 2023 revealed that Adult Genetics handled the majority (75.7%), followed by Pediatric Genetics (17.4%) and Adult Cancer Risk Genetics (6.9%) [TABLE 1]. While 61.1% of these consults were completed electronically, 31.9% were declined and converted to in-person visits [TABLE 3], suggesting e-Consults can help determine when face-to-face evaluations are needed. Nearly half (47.9%) of FY 2023 e-Consults came from generalists in primary care, general medicine, and women’s health [TABLE 1], indicating broad adoption across non-genetics specialties. e-Consultants provided prompt responses, spending 20 minutes or less on 75% of cases [TABLE 4].

The most common reasons for e-Consults were questions about genetic diagnoses (50.6%), test interpretation (11.1%), and patient management [TABLE 5]. e-Consultants frequently recommended clinic visits (42.3%), provided counseling and management advice (24.3%), and offered guidance on genetic testing (11.8%) [TABLE 6]. Referral outcomes data showed that e-Consults led to 64% of patients being seen or scheduled with genetics, while 20% had their care informed by e-Consult advice without a formal Genetics referral [FIGURE 1].

These findings align with and expand upon the limited existing literature on Genetics e-Consult programs. Similar to studies by Bhola et al.^9^ and Folkerts et al.^10^, we observed efficient turnaround times, a wide range of clinical topics addressed, and high rates of guideline-concordant care when e-Consults provided actionable recommendations. Our higher e-Consult volume likely reflects the longer duration and wider scope of our program.

The high proportion of e-Consults completed electronically (57.8%) highlights the potential for e-Consults to optimize genetics care delivery by addressing straightforward questions, triaging complex cases for in-person evaluation, and guiding appropriate pre-visit workup. The 84% adherence rate among providers to e-Consult recommendations underscores the educational impact of specialist guidance.

Limitations of our study include the single-center design, lack of long-term outcomes data, and absence of referring provider and patient perspectives. Future research should compare e-Consult programs across diverse healthcare settings, incorporate feedback from key stakeholders, and evaluate the impact on care access, quality, and costs.

In conclusion, the UCSF Genetics e-Consult service demonstrates a scalable and efficient model for expanding access to genetics expertise. By providing timely, individualized guidance to non-genetics providers, e-Consults can help optimize resource utilization, provider education, and patient care as genetics becomes increasingly integrated into routine clinical practice.

## Data Availability

All data produced in the present study are available upon reasonable request to the authors

